# Phase II trial of chemotherapy, cetuximab and erlotinib in patients with metastatic or recurrent squamous cell carcinoma of the head and neck

**DOI:** 10.1101/2025.01.07.25320123

**Authors:** Aarti Bhatia, Ranee Mehra, Jessica Bauman, Saad A. Khan, Wei Wei, Veronique Neumeister, Teresa Sandoval-Schaefer, R. Katherine Alpaugh, Miriam Lango, David L. Rimm, John A. Ridge, Barbara Burtness

## Abstract

Prognosis for patients with recurrent/metastatic (R/M) head and neck squamous cell cancer (HNSCC) remains poor. We hypothesized that the addition of an epidermal growth factor receptor (EGFR) tyrosine kinase inhibitor (TKI) to standard therapy would improve responses by inhibiting nuclear translocation of EGFR and designed a phase 2 trial of chemotherapy, cetuximab and erlotinib in patients with R/M HNSCC.

24 patients were enrolled and treated with carboplatin, paclitaxel and cetuximab administered in 21-day cycles. Erlotinib was added with cycle 2. Primary end point was objective response rate (ORR). Secondary end points were toxicity, overall survival (OS) and laboratory correlates.

Median age was 65.5 years. Median duration on treatment was 4.6 months. ORR with cycle 1 of treatment was 33.3%, and for cycle 2 and beyond was 62.5%. Median progression-free survival (PFS) was 6.2 months, and median OS was 10.6 months. Most common treatment-related adverse events included anemia, neutropenia, skin rash, diarrhea, and hypomagnesemia.

Dual EGFR blockade was tolerable and efficacious in this small patient sample. With an ORR of 62.5%, the study met its primary endpoint. PFS and OS were comparable to historical controls. Dual EGFR targeting without the chemotherapy backbone is worthy of further study.

Clinical trial information: NCT01316757. Study registered March 8^th^, 2011

## Introduction

### Epidemiology

Squamous cell cancers of the head and neck (HNSCC) are a heterogenous group of morbid cancers most frequently caused by longstanding exposure to tobacco and alcohol or by the Human Papillomavirus (HPV) (1). More than 700,000 patients around the world were afflicted in 2022 and HNSCC ranks among the top ten most frequently diagnosed cancers in men in the United States (2, 3). Although 90% of patients present with local or locoregional disease, curative intent treatment is usually multi-modal and disfiguring, despite which up to 40% of patients go on to develop a local recurrence or distant disease (1). Treatment options for unresectable recurrent or distant metastatic disease usually include palliative radiation or systemic therapy. First-line FDA-approved systemic therapy is the anti-programmed death-1 (anti-PD-1) immune checkpoint inhibitor pembrolizumab either as a single agent or in combination with platinum and 5-fluorouracil chemotherapy (4). Subsequent lines of systemic therapy include other chemotherapeutic agents like taxanes or methotrexate and the anti-epidermal growth factor receptor (EGFR) monoclonal antibody cetuximab, which leads to single agent response rates of approximately 11-13% (5). Despite recent advances in systemic therapy and our greater understanding of oncogenic drivers and mechanisms of drug resistance, median overall survival for patients diagnosed with recurrent or metastatic (R/M) disease remains dismal and there is an urgent unmet need for novel therapies and drug combinations.

### Targeting EGFR in HNSCC

EGFR is a transmembrane glycoprotein that is overexpressed in more than 80% of HNSCC and is associated with a worse clinical outcome, making it a rational therapeutic target (6). Cetuximab is an immunoglobulin G1 (IgG1) monoclonal antibody targeting the extracellular domain of EGFR and has a dual mechanism of action: it blocks ligand-stimulated EGFR signaling and fosters EGFR internalization. In preclinical models cetuximab had also been shown to stimulate antibody-dependent cell-mediated cytotoxicity (ADCC) against EGFR-expressing tumor cells (7, 8). Cetuximab is well-tolerated alone or in combination with cisplatin (9, 10). Based on improved outcomes demonstrated in two phase 3 trials, cetuximab was FDA-approved in 2006 in combination with radiation therapy (RT) for the treatment of locally advanced HNSCC (11) and in 2011 in combination with platinum and 5-fluorouracial for the treatment of R/M HNSCC (12). To date, it remains the only FDA approved targeted therapy in this disease.

Cetuximab activity is limited however, by intrinsic and acquired mechanisms of resistance. These include: hetero-dimerization with other ErbB family members and downstream signaling (13), alterations in downstream signaling pathway proteins like HRAS or PI3K (14, 15), compensatory activation via crosstalk with other receptor tyrosine kinases (RTKs) such as ErbB2, ErbB3 and MET (16–19), tumor hypoxia mediated activation of downstream receptors leading to increased tumor cell migration (20) and immune suppressive signaling mechanisms, e.g. via high TGF-β expression and inhibition of natural killer (NK) cell populations, that enable tumor cells to bypass the ADCC effect of cetuximab (21, 22). We have previously demonstrated nuclear translocation of the EGF receptor, where it activates cyclin D1 and stabilizes proliferating cell nuclear antigen (PCNA), thus enhancing tumor proliferation and angiogenesis (23, 24) and possibly serving as a source of cetuximab resistance as well.

### Study Rationale

Membrane expression of EGFR correlates with a poorer prognosis in several solid tumor types, such as head and neck and esophageal carcinomas. Moreover, the phosphorylated or active form of EGFR (pEGFR) can translocate to the nucleus, where it serves as a co-transcription factor for critical processes like cellular proliferation and angiogenesis and is highly associated with worse survival outcomes, disease progression and treatment resistance (23, 25). EGFR tyrosine kinase inhibitors (TKIs) are oral drugs that competitively bind with the intracellular tyrosine kinase domain of EGFR to inhibit downstream signaling (26) and may inhibit signaling from EGFR in a ligand-independent manner. Several agents have undergone testing in HNSCC with modest monotherapy activity given the lack of EGFR activating mutations in this disease (27–29). However, preclinical models suggest synergy between antibody and TKI EGFR targeting in HNSCC cell lines, leading to a 3-to-4-fold enhancement of apoptosis (30, 31). Lapatinib, a dual inhibitor of EGFR and HER-2, has been shown in cell lines to reduce levels of nuclear EGFR and HER-2, possibly by inhibiting translocation of these receptors to the nucleus (32). Phase I data from Johns Hopkins had demonstrated that an EGFR targeting antibody and TKI could be safely co-administered in humans [personal communication, A. Forastiere] and a phase I trial of erlotinib with cetuximab and bevacizumab supported the conception of this study (33). The current study therefore hypothesized that dual targeting of EGFR with cetuximab and a TKI would overcome these postulated cetuximab resistance mechanisms, inhibiting both ligand dependent and independent signaling of the pathway and preventing nuclear effects of EGFR via intracellular activity of the TKI. We hence conducted a single-arm phase II trial evaluating the efficacy of the TKI erlotinib when added to standard chemotherapy and cetuximab in patients with treatment-naïve R/M HNSCC. Tissue was obtained for correlative analysis.

## Materials and Methods

### Patients

This multi-center study was conducted at Yale Cancer Center (New Haven, CT), Fox Chase Cancer Center (Philadelphia, PA) and at the University of Texas Southwestern Cancer Center (Dallas, TX). The study protocol was approved by all local institutional review boards, performed in accordance with Good Clinical Practice, and registered with ClinicalTrials.gov (identifier NCT01316757). Written informed consent was provided by all study participants. Primary inclusion criteria included the following: histologically confirmed, treatment-naïve, R/M squamous cell cancer of any primary mucosal site in the head and neck, an Eastern Cooperative Oncology Group performance status (ECOG PS) score of 0-1, measurable disease based on RECIST version 1.1, adequate organ and marrow function and willingness to provide baseline and on-treatment tumor biopsies if safe and medically feasible. Prior chemotherapy or cetuximab were permitted as part of multi-modality treatment of locally advanced disease if completed at least 4 months prior to study enrollment. No prior EGFR TKI, investigational EGFR or panErbB reversible or irreversible inhibitor or any prior panitumumab or investigational EGFR-directed monoclonal antibody was permitted.

### Study Design

This was a single-arm, open label, phase 2 study designed to evaluate the efficacy of erlotinib when added to combination carboplatin/paclitaxel/cetuximab systemic therapy in patients with metastatic or recurrent HNSCC, who had not previously received systemic therapy for this stage of their cancer. This study was designed and enrolled patients prior to the approval of anti-programmed death-1 (anti-PD-1) agents in HNSCC. The study schema is depicted in Figure 1. Planned enrollment was 43 patients. Baseline tumor assessments and biopsy were performed, after which cycle 1 treatment was initiated with carboplatin/paclitaxel/cetuximab systemic therapy. Following a 21-day cycle, tumor assessment and biopsy were repeated, followed by resumption of therapy with the addition of oral erlotinib and a subsequent biopsy. The primary endpoint was objective response rate (ORR, complete plus partial response, CR+PR) as determined by RECIST version1.1. The ORR was to be compared to historical controls in the pre-immunotherapy era (12); each patient also served as a self-control between cycles 1 and 2. Key secondary endpoints were toxicity, overall survival (OS), and laboratory correlates to determine whether EGFR signaling was more effectively inhibited after the addition of erlotinib than it was after chemotherapy/cetuximab alone. Adverse events (AEs) were graded with the National Cancer Institute Common Terminology Criteria for Adverse Events, version 4.0 with the exception of skin rash which was graded as tolerable (Grade 1 to 2) or intolerable (Grade 3 to 4).

**Figure 1.**
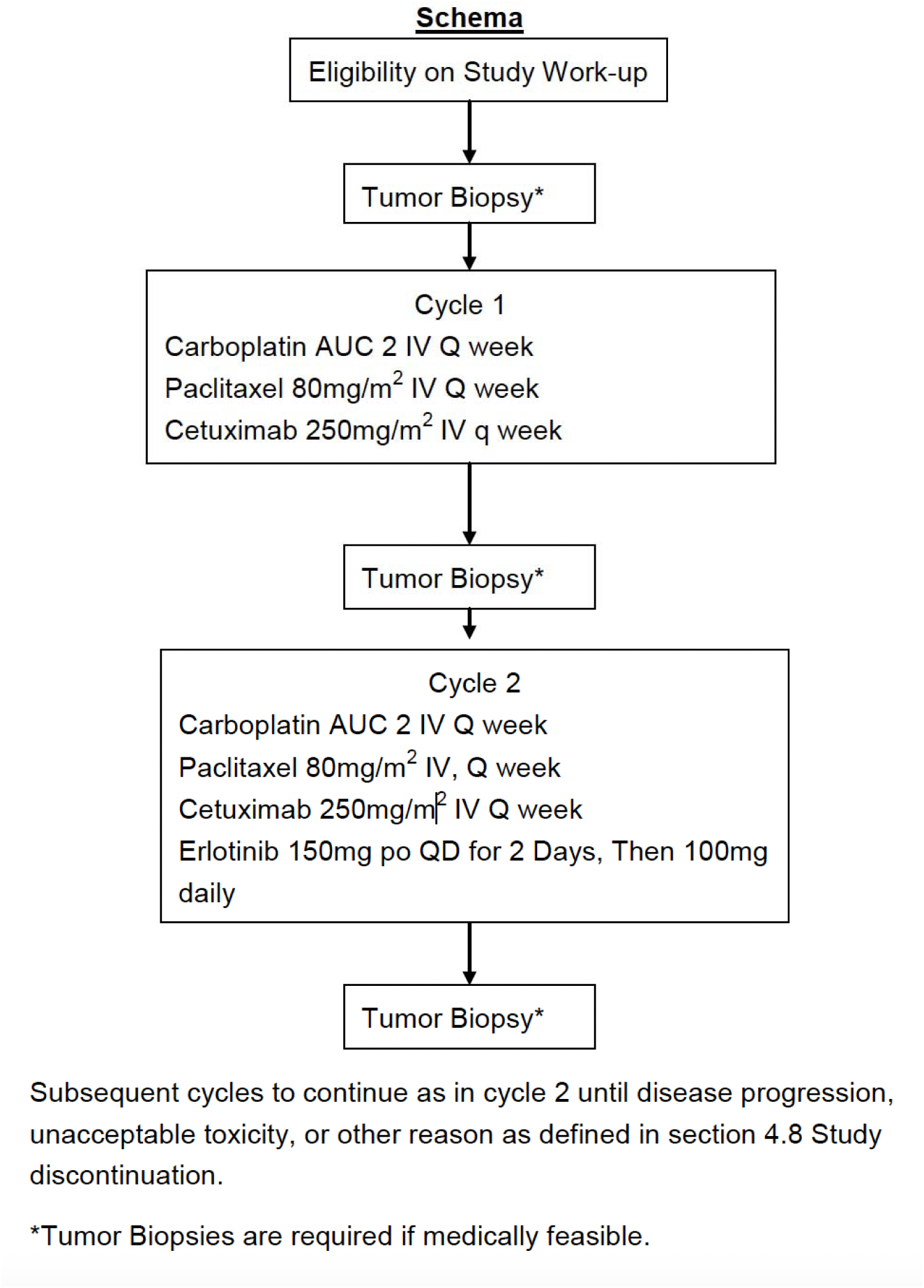
Study Schema.

### Treatment Plan

Each treatment cycle consisted of three weeks. Drug doses were calculated on day one of each cycle. Agents were administered in the following order: cetuximab followed by paclitaxel and then carboplatin. Erlotinib was self-administered by the patient. Following a 400mg/m^2^ loading dose, cetuximab was administered at 250 mg/m^2^ intravenously (IV) every week, paclitaxel at 80 mg/m^2^ IV every week, and carboplatin at a projected area under the curve (AUC) of 2 administered IV every week. Erlotinib was supplied in tamper-proof bottles containing 30 pills and was self-administered continuously beginning with day 1 of cycle 2. Erlotinib was dosed at 150 mg orally daily for two days followed by 100 mg orally daily. Erlotinib was to be taken at the same time each day with 200 mL of water, one hour before or two hours after a meal. Patients unable to swallow tablets could dissolve them in distilled water for administration via a percutaneous endoscopic gastrostomy (PEG) tube. Pre-medications to decrease the risk of hypersensitivity were required prior to the first dose of cetuximab and of paclitaxel, modifications thereafter were permitted per the treating physician if no reactions were noted. In the event of serious or intolerable AEs, two levels of dose reduction were available for cetuximab (200 mg/m^2^ and 150 mg/m^2^ IV every week), paclitaxel (70 mg/m^2^ and 60 mg/m^2^ IV every week), carboplatin (AUC 1.5 and AUC 1 weekly) and erlotinib (50 mg and 25 mg orally daily). Dose modifications were undertaken on a drug-specific basis. Dose re-escalation was not permitted and missed doses were not made up. Tumor assessments were performed using physical examination and contrast-enhanced computed tomography (CT) or magnetic resonance imaging (MRI) scans at baseline, at the beginning of cycle 2, at the beginning of cycle 4, and every nine weeks thereafter. Tumor biopsy was performed prior to the start of cycle 1, after cycle 1 and after cycle 2 when safe and medically feasible. Protocol treatment was administered until progression, unacceptable toxicity, or withdrawal of consent. The study schema is depicted in Figure 1.

### Biomarker Analyses

Tissue microarrays were constructed from large bore core biopsies or excisional specimens obtained from enrolled patients, chromogenic and fluorescent immunohistochemical (IHC) assays were performed and analyzed by pathologist read-out respectively with automated quantitative image analysis (AQUA, Navigate Biopharma, Carlsbad, CA, (34)) of fluorescently labeled antigens. Following biomarkers were analyzed: EGFR, phospho-EGFR, nuclear EGFR, nuclear pEGFR, Her-2, phospho-Her-2, Her-3, Her-4, FGF, phospho-Akt, PTEN and Caspase 3. Because of the limitations imposed by the trial size, correlation of IHC results with response and survival data was exploratory only.

### Statistical Analyses

The primary endpoint of ORR was determined on protocol-specified tumor assessments and assessed by RECIST version 1.1. Exact binomial inference was used to investigate whether study objectives were met. Kaplan-Meier curves were used to describe overall survival. Wilcoxon paired-sample test was used to investigate whether EGFR assay levels in patients varied between cycle 1 (before erlotinib was added) and cycle 2 (after erlotinib was added). Similar tests were performed for the biomarkers related to EGFR. Spearman correlations were used to assess the associations of the biomarkers with each other.

The null hypothesis was an ORR of 21% and the alternative was 40%. A death or loss to follow up without documented and confirmed response would be considered a response failure. The study would be considered a success if thirteen or more (30% or more) of the participants had responses. A two-stage design was used for the study to stop for futility. We examined the data after the first fifteen patients were accrued and study was to proceed only if three or more of the first fifteen had objective responses. With a total of 43 patients, the Type I error rate for this study, accounting for the early stopping rule for futility, was 9.5%. The power was 91.5%. The Type I error rate was slightly above the traditional level of 5% for reasons elaborated by Rogatko and Litwin (1996).

## Results

### Patient Characteristics

Twenty-four patients were enrolled between March 2011 and October 2014, of whom twenty-three initiated protocol treatment. One patient did not begin treatment because of failure to attend the protocol-required treatment appointments. The study was terminated due to slow accrual. Baseline characteristics of the study population were typical for this disease and are outlined in Table 1. Median age was 65.5 years (range 40 – 82 years) and fifteen of twenty-four (63%) of enrolled patients were male. Three of twenty-four (12%) were under-represented racial or ethnic minorities. Oropharyngeal primaries represented a small proportion of the enrolled patient population (6/24, 25%) and p16 status was available on only two of those six patients. Efficacy outcomes were therefore not analyzed by p16 status given lack of power.

**Table 1:** Baseline Characteristics.

|  |  |  |  |
| --- | --- | --- | --- |
| **Gender** | Male | 15/24 | 63 |
|  | Female | 9/24 | 37 |
| **ECOG PS** | 0 | 2/24 | 8 |
|  | 1 | 22/24 | 92 |
| **Primary Tumor Site** | Oropharynx | 6/24 | 25 |
|  | Larynx | 4/24 | 17 |
|  | Lip/Oral Cavity | 6/24 | 25 |
|  | Hypopharynx | 2/24 | 8 |
|  | Unknown | 6/24 | 25 |
| **Race** | Caucasian | 18/24 | 75 |
|  | African-American | 2/24 | 8 |
|  | Asian | 3/24 | 13 |
|  | Hispanic | 1/24 | 4 |

### Protocol Treatment

At the data cutoff date of March 2019, all patients had discontinued treatment. Disease progression was the predominant reason for treatment discontinuation, accounting for fifteen of twenty-four (62.5%) patients. Five patients were alive at the time of data cutoff. The median number of treatment cycles was 6 (range 0 – 17). Median duration on treatment was 4.6 months (range 0 – 12.9 months). One patient discontinued protocol therapy after cycle 1, day 1 due to grade 4 hypersensitivity reaction to cetuximab. Four patients withdrew consent for treatment – one because they had achieved complete response to treatment and went on to receive curative intent radiation, one for treatment-related nausea and fatigue after cycle 1, day 1 of treatment, the third patient also withdrew due to treatment-related fatigue following 6 cycles of treatment and the fourth withdrew for treatment-related neutropenia after 3 cycles of treatment. The last patient achieved partial treatment response (PR) which was maintained at 17.9 months, which was the date of last contact. One patient discontinued treatment after 4 cycles on study due to a planned vacation and exceeding the maximum allowable time between treatment cycles.

### Efficacy

The primary endpoint of response rate (ORR) was assessed for twenty of twenty-four patients with cycle 1 of treatment (prior to erlotinib treatment being initiated) and again post cycle 2 after erlotinib treatment began. Four patients who were enrolled but did not complete a cycle of treatment, did not have response assessed and were considered response failures. ORR with cycle 1 of treatment was 33.3% (95% CI: 15.6%-55.3%), including 7 partial responses (PR) and 1 complete response (CR). ORR for cycle 2 and beyond was 62.5% (95% CI: 40.6%-81.2%), including 13 PR and 2 CR. Patients who achieved CR remained cancer-free 12- and 6-years after initiating trial treatment respectively. The ORR met protocol-specified criteria for treatment success. Disease control rate (DCR) for cycle 1 and for cycle 2 and beyond was 83.3%, with 12 patients (50%) achieving stable disease post cycle 1 and 5 patients (20.8%) achieving stable disease after cycle 2. ORR for study participants is outlined in Table 2.

**Table 2:**
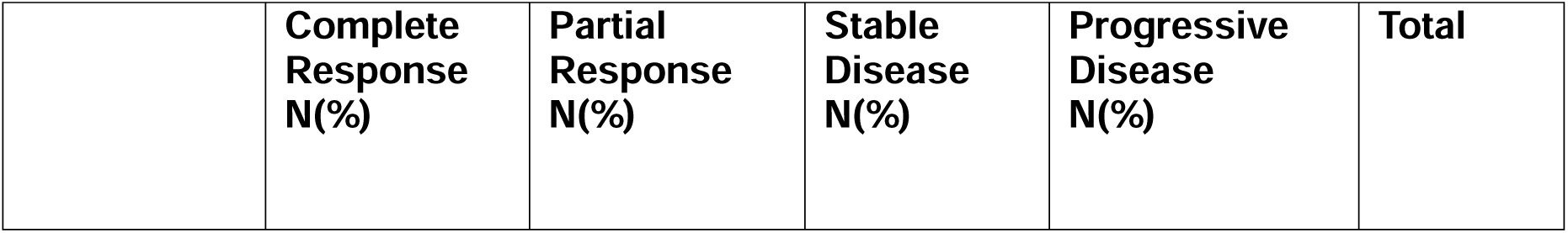
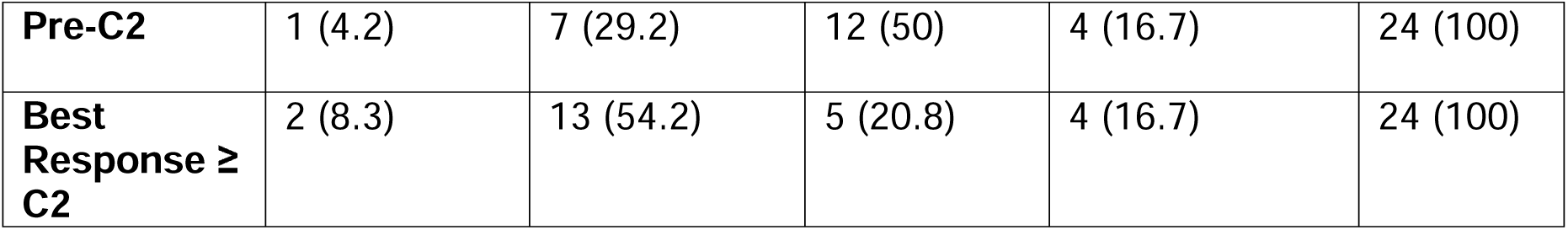
Response Assessments.

|  | **Complete Response**<br>**N(%)** | **Partial Response**<br>**N(%)** | **Stable Disease**<br>**N(%)** | **Progressive Disease**<br>**N(%)** | **Total** |
| --- | --- | --- | --- | --- | --- |
| **Pre-C2** | 1 (4.2) | 7 (29.2) | 12 (50) | 4 (16.7) | 24 (100) |
| **Best Response ≥ C2** | 2 (8.3) | 13 (54.2) | 5 (20.8) | 4 (16.7) | 24 (100) |

Median progression-free survival (PFS) was 6.2 months (95% CI: 5.2-8.5) and median overall survival was 10.6 months (95% CI: 6.9-32.5) based on 23 patients in the intent-to-treat population. Among patients who completed at least a cycle of therapy, 12 months OS was 50%- and 18-months OS was 25%. PFS and OS Kaplan-Meier (KM) curves for the population are shown in Figure 2.

**Figure 2:**
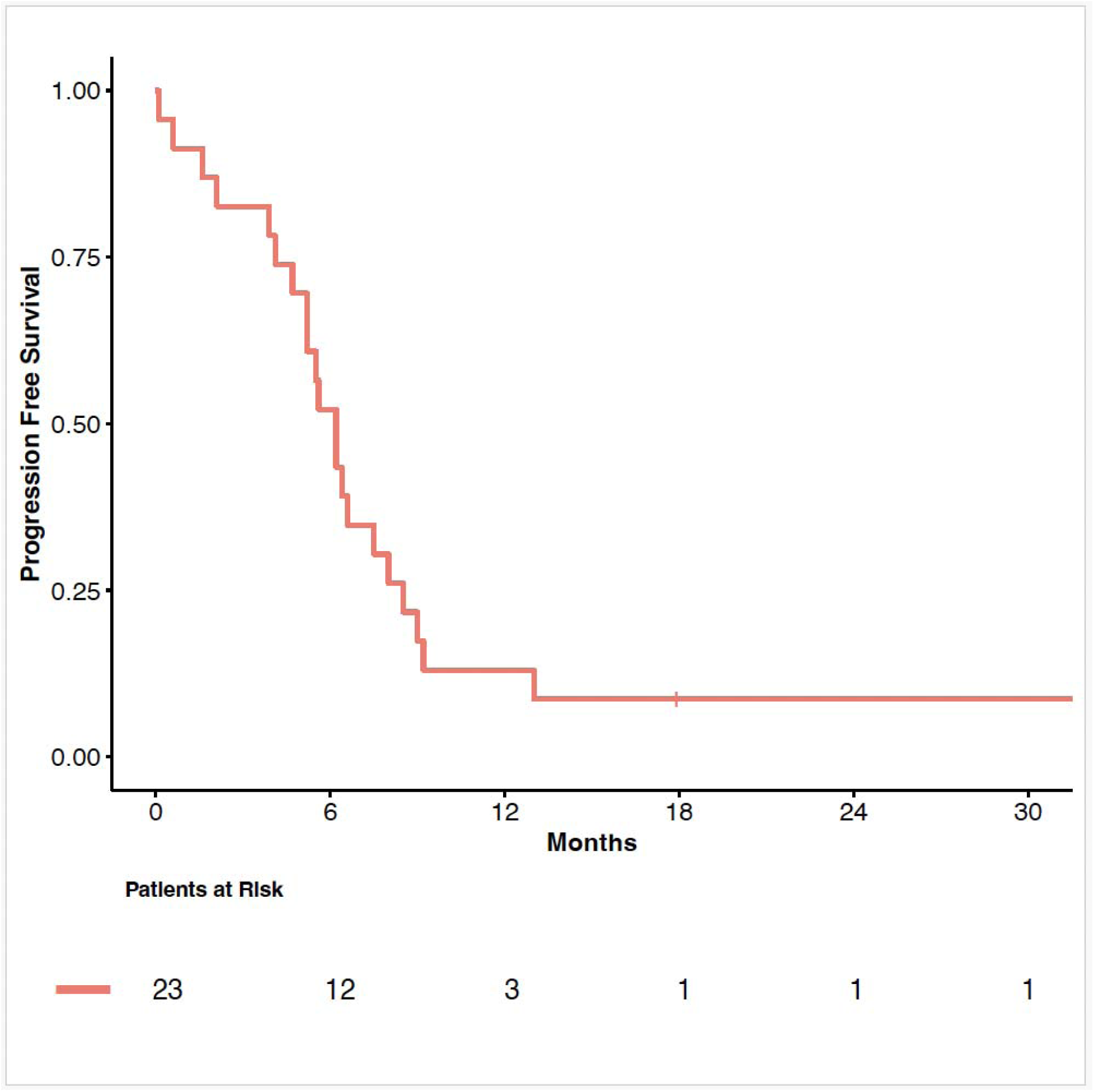
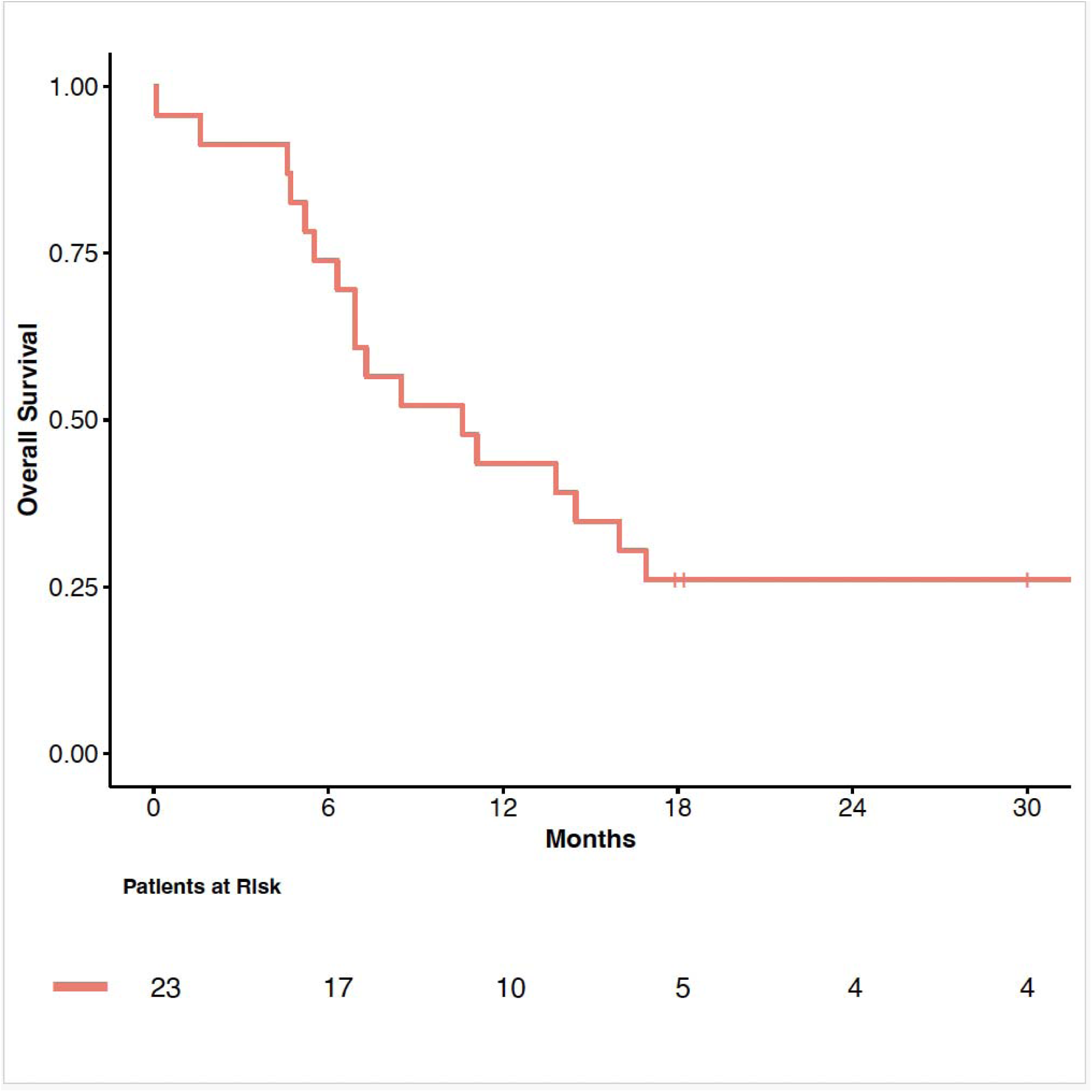
PFS and OS KM Curves.

### Toxicity

All treatment-emergent adverse events, regardless of attribution, are summarized in Table 3. Most common treatment-related adverse events, occurring in over 50% of patients, included anemia, neutropenia, skin rash, diarrhea, and hypomagnesemia. The last three are class toxicities associated with EGFR inhibition. One patient developed fatal hypoxic respiratory failure due to pneumonitis attributed to erlotinib after 7 cycles of trial treatment. Pneumonitis is a rare but known class toxicity effect of EGFR inhibitors (35). Another patient died of a carotid bleeding event attributed to study treatment after 3 cycles of protocol therapy. A third patient received cycle 1, day 1 of treatment and was found dead at home two days later. An autopsy was not required, and death was attributed not related to study treatment by the treating investigator.

**Table 3:** Common Adverse Events.

| **ORGAN SYSTEM** | **GRADE 1 – 2** | **GRADE ≥ 3** | **TOTAL N/24 (%)** |
| --- | --- | --- | --- |
| Anemia | 11 | 7 | 18/24 (75%) |
| Neutropenia | 10 | 8 | 18/24 (75%) |
| Leukopenia | 7 | 10 | 17/24 (71%) |
| Sensory Neuropathy | 5 | 3 | 8/24 (33%) |
| Fatigue | 17 | 1 | 18/24 (75%) |
| Alopecia | 7 | 0 | 7/24 (29%) |
| Rash | 12 | 5 | 17/24 (71%) |
| Nausea | 10 | 0 | 10/24 (42%) |
| Diarrhea | 15 | 3 | 18/24 (75%) |
| Constipation | 12 | 0 | 12/24 (50%) |
| Hypomagnesemia | 10 | 5 | 15/24 (63%) |
| Hypoalbuminemia | 15 | 0 | 15/24 (63%) |
| Increased AST/ALT | 12 | 0 | 12/24 (50%) |
| Mucositis | 7 | 1 | 8/24 (33%) |

### Biomarkers

Tumor biopsies were collected at a minimum of two or more time points for eight patients enrolled on this study. Changes in EGFR expression and PCNA with trial treatment were analyzed for these patients (Figure 3). Treatment response to cetuximab and PFS were also stratified by baseline PTEN status. In view of the small numbers, this analysis was exploratory only.

**Figure 3:**
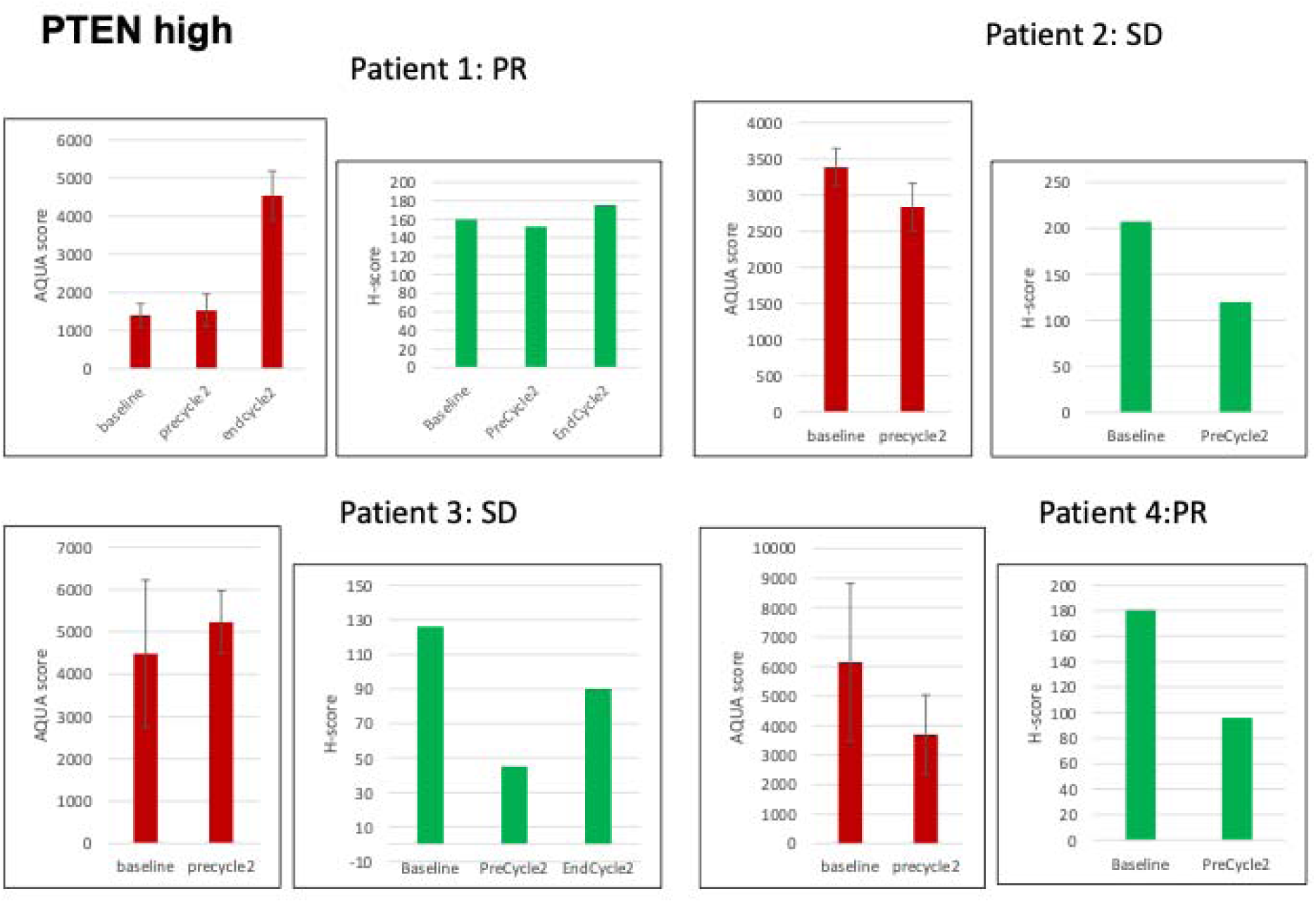
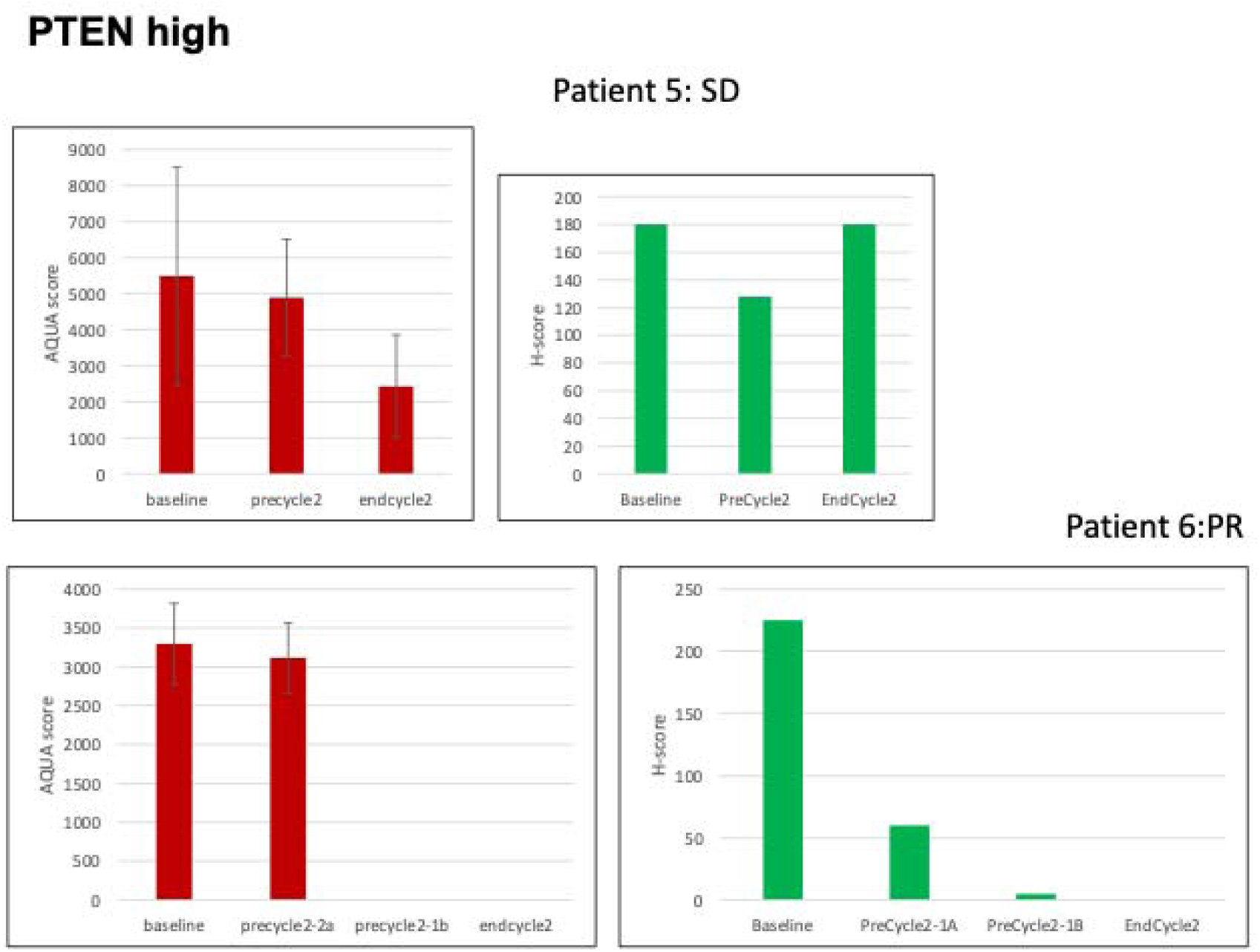
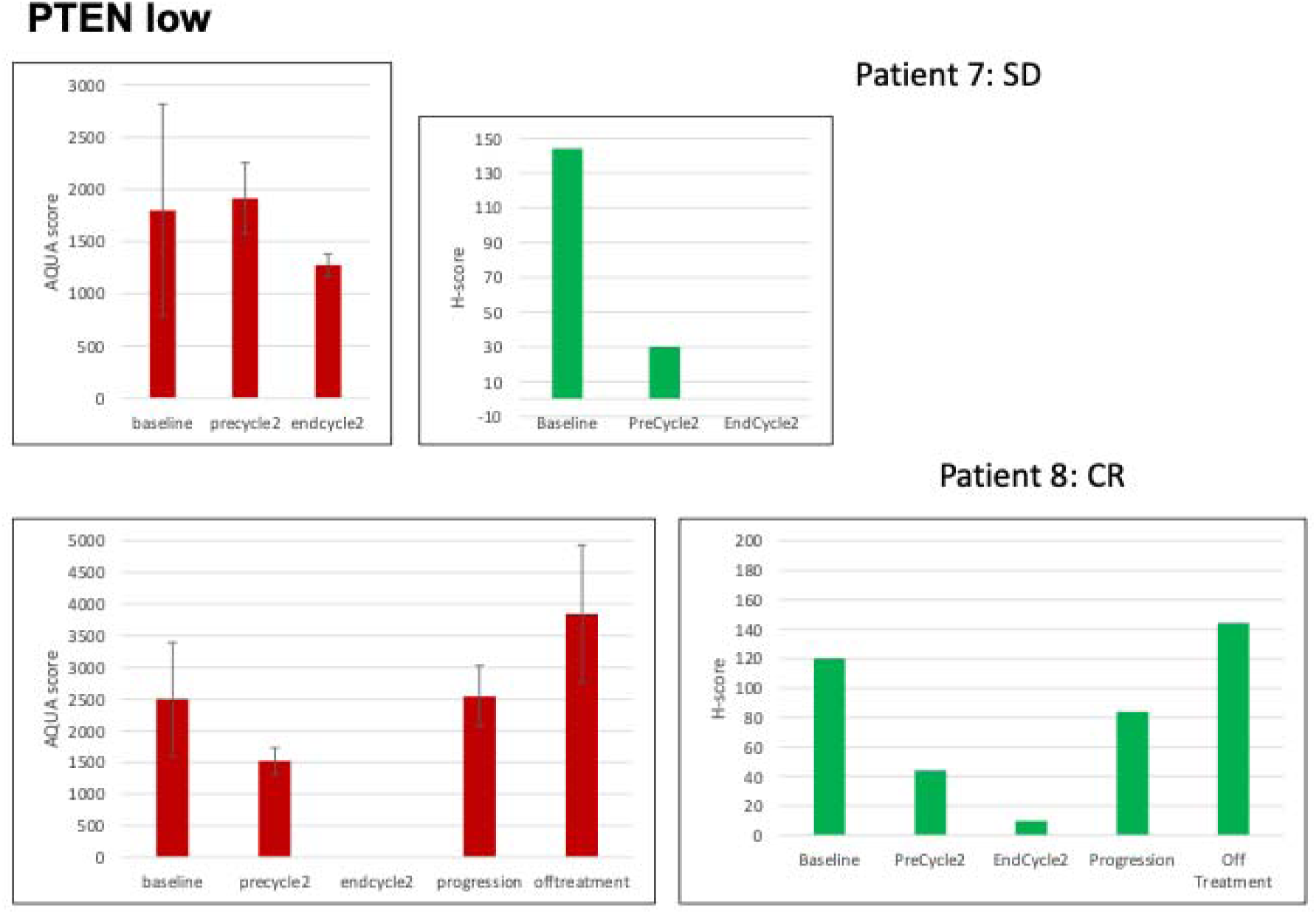
Changes in EGFR and PCNA expression with trial treatment. Red bars represent EGFR expression (analyzed using the AQUA method and reported as AQUA scores) and green bars represent PCNA expression (evaluated by chromogenic IHC and reported as H-scores). Patients are sorted by PTEN low or high status.

Cetuximab reduced EGFR expression in 5 of 8 patients with paired specimens, and the addition of erlotinib further reduced EGFR expression in 2 of 3 patients with paired post-cetuximab and post-cetuximab plus erlotinib specimens. The EGFR AQUA scores decreased by an average of 22% after treatment with cetuximab and by a further 42% with the subsequent addition of erlotinib. Erlotinib added to cetuximab reduced PCNA relative to cetuximab treatment in 2 of 8 patients, including in a PTEN-low patient. The AQUA score was reduced by an average of 85% with the addition of erlotinib. 3 of 6 patients with PTEN-high tumors and 1 of 2 patients with PTEN-low tumors had objective tumor response to cycle 1 treatment with cetuximab. Median PFS was numerically slightly longer for patients with PTEN-high tumors (7.7 months) when compared to the overall enrolled population.

## Discussion

This study is a single-arm, open-label, phase 2 trial evaluating the efficacy of a platinum/taxane chemotherapy backbone with dual EGFR inhibition using the mAb cetuximab and the small molecule TKI erlotinib in the first-line treatment of patients with recurrent or metastatic HNSCC. Comparison of efficacy outcomes was made to a historical control of the EXTREME trial (12), the standard of care at the time this trial was conducted, which tested a platinum/5-fluorouracil combination with cetuximab in a similar patient population. The ORR of 62.5% with chemotherapy and dual EGFR inhibition exceeded the ORR of 36% seen with the triple drug combination in the EXTREME trial and in the more recent pembrolizumab plus chemotherapy (ORR 36%) and cetuximab plus chemotherapy (ORR 36%) groups of the randomized Keynote 048 trial (4). This study therefore met its primary endpoint. Remarkably, one of two patients who achieved radiologic CR with study treatment, was able to be consolidated with curative intent therapy. This patient withdrew from the study after achieving radiologic PR to pursue surgical treatment and was found to have a pathologic CR at the time of resection. She was alive and cancer-free 12 years after trial enrollment. The second patient who achieved CR also remains cancer-free at date of last contact, 6 years since initiation of trial treatment. Median PFS and OS for the entire cohort were comparable to those reported in the EXTREME trial. However, therapy with dual EGFR targeting added to chemotherapy proved toxic, with nearly three quarters of patients experiencing fatigue, cytopenias and skin rash. 4 of 24 patients who enrolled on this study did not complete the first cycle of study treatment, which might at least partly be explained by frailer patients being selected for a carboplatin- and paclitaxel-based regimen in an era where cisplatin and 5-flurouracil was the standard of care, than the toxicity of the platinum doublet and cetuximab combination.

Biomarker analysis of serial samples in a subset of patients demonstrated that erlotinib plus cetuximab decreased EGFR expression and reduced PCNA levels, as a surrogate marker of the non-canonical nuclear actions of EGFR, more effectively than did cetuximab alone. These findings demonstrate the potential to achieve more effective EGFR inhibition with dual antibody and kinase inhibitor regimens, and the need to assess nuclear EGFR levels and activity when evaluating EGFR-targeted therapies. Recognizing the remarkable tumor activity seen with dual EGFR blockade in this trial, we subsequently designed another single-arm phase 2 study exploring the combination of cetuximab and the irreversible, pan-EGFR TKI afatinib in patients with platinum- and/or anti-PD-1-refractory, recurrent or metastatic HNSCC (NCT02979977). This trial is nearing completion of enrollment.

With the more recent advent of anti-PD-1 therapy in HNSCC, ongoing clinical research in this disease is broadly focused on two distinct paradigms of systemic treatment – improving responses to immune checkpoint inhibition and overcoming resistance to EGFR inhibition. Multiple recent clinical trials have demonstrated superior ORR and PFS for dual targeting of EGFR and other pathways it engages in crosstalk with, compared to the historical responses reported with cetuximab monotherapy (36, 37). This underscores the ongoing relevance of EGFR-directed therapy in immunotherapy-experienced patients with HNSCC.

Multiple other novel therapeutics including bi-specific antibodies, antibody-drug conjugates and combinations have demonstrated a preclinical signal of activity or are in clinical development for early- and late-phase trials in HNSCC. The combination of cetuximab and ficlatuzumab will be examined in a randomized phase 3 trial in patients with pan-refractory R/M HNSCC (NCT06064877). The bifunctional EGFR/TGFβ inhibitor BCA101, in combination with pembrolizumab, was tested in the first-line treatment of a R/M HNSCC expansion cohort of a phase 1/1b trial and results from 31 patients were reported at the American Society of Clinical Oncology (ASCO) meeting in June 2023 (38). 65% ORR was reported in the HPV-negative cohort and the combination will be evaluated in a randomized study in this patient population. Amivantamab, an IgG1 EGFR-MET–bispecific mAb with US FDA approval for EGFR Exon 20 insertion–driven lung cancer, was administered in combination with pembrolizumab into EGFR and MET-expressing tumor-bearing patient-derived xenograft (PDX) models (39). The combination could reduce the EGFR^high^MET^high^ cluster, which enabled immune evasion, and created a favorable immune tumor microenvironment that could effectively control tumor growth. A phase 1 trial is currently enrolling patients with HNSCC (NCT06385080).

This trial demonstrated that chemotherapy combined with cetuximab and erlotinib leads to a high response rate, including a durable CR, and that dual EGFR targeting impacts EGFR and PCNA expression. The generalizability of these findings is limited by the small sample size of this study, as well as the toxicity of the regimen. However, the remarkable activity seen here adds to a growing body of evidence supporting the further testing of EGFR inhibition-based strategies in HNSCC, especially in the immune checkpoint-inhibitor experienced HPV-negative subset, where prognosis is poor and there remains a critical, unmet need for novel therapeutics.

## Acknowledgements

Genentech supplied Erlotinib to all patients enrolled on this trial. Results from this trial were presented in Poster format at the 2016 ASCO Annual Meeting: <u>Aarti K. Bhatia et al.</u>, Phase II trial of carboplatin/paclitaxel and cetuximab, followed by carboplatin/paclitaxel/cetuximab and erlotinib, in metastatic or recurrent squamous cell carcinoma of the head and neck.. *JCO* **34**, 6027-6027(2016). DOI:<u>10.1200/JCO.2016.34.15_suppl.6027</u>

## Authors’ Contributions

All authors conceived and/or designed the work that led to the submission, acquired data, and/or played an important role in interpreting the results, drafted or revised the manuscript, approved the final version and agreed to be accountable for all aspects of the work in ensuring that questions related to the accuracy or integrity of any part of the work are appropriately investigated and resolved.

## Ethics Approval and Consent to Participate

The study was conducted according to the International Conference on Harmonization E6 Guideline for Good Clinical Practice (GCP) and the Declaration of Helsinki. IRBs of all participating institutions (Yale Cancer Center, Fox Chase Cancer Center and University of Texas Southwestern) approved the study. All enrolled patients signed the IRB approved informed consent before study assessments were performed and treatment was administered.

## Data Availability Statement

The data generated in the present study are included in the figures and/or tables of this article.

**Patient Consent was obtained for publication of study findings.**

---

|  | **N/24** | **%** |
| --- | --- | --- |
| **Age (Median [Min, Max])** | 65.5 years (40 years, 82 years) |  |

## References

1. Chow LQM. Head and Neck Cancer. N Engl J Med. 2020;382(1):60–72.

2. Siegel RL, Miller KD, Wagle NS, Jemal A. Cancer statistics, 2023. CA Cancer J Clin. 2023;73(1):17-48.

3. Bray F, Laversanne M, Sung H, Ferlay J, Siegel RL, Soerjomataram I, Jemal A. Global cancer statistics 2022: GLOBOCAN estimates of incidence and mortality worldwide for 36 cancers in 185 countries. CA Cancer J Clin. 2024;74(3):229-63.

4. Burtness B, Harrington KJ, Greil R, Soulieres D, Tahara M, de Castro G, Jr., et al. Pembrolizumab alone or with chemotherapy versus cetuximab with chemotherapy for recurrent or metastatic squamous cell carcinoma of the head and neck (KEYNOTE-048): a randomised, open-label, phase 3 study. Lancet. 2019;394(10212):1915–28.

5. Bhatia A. Targeting Epidermal Growth Factor Receptor in Head and Neck Cancer. Cancer J. 2022;28(5):331–8.

6. Nicholson RI, Gee JM, Harper ME. EGFR and cancer prognosis. Eur J Cancer. 2001;37 Suppl 4:S9–15.

7. Kimura H, Sakai K, Arao T, Shimoyama T, Tamura T, Nishio K. Antibody-dependent cellular cytotoxicity of cetuximab against tumor cells with wild-type or mutant epidermal growth factor receptor. Cancer Sci. 2007;98(8):1275–80.

8. Li S, Schmitz KR, Jeffrey PD, Wiltzius JJ, Kussie P, Ferguson KM. Structural basis for inhibition of the epidermal growth factor receptor by cetuximab. Cancer Cell. 2005;7(4):301–11.

9. Baselga J, Pfister D, Cooper MR, Cohen R, Burtness B, Bos M, et al. Phase I studies of anti-epidermal growth factor receptor chimeric antibody C225 alone and in combination with cisplatin. J Clin Oncol. 2000;18(4):904–14.

10. Burtness B, Goldwasser MA, Flood W, Mattar B, Forastiere AA, Eastern Cooperative Oncology G. Phase III randomized trial of cisplatin plus placebo compared with cisplatin plus cetuximab in metastatic/recurrent head and neck cancer: an Eastern Cooperative Oncology Group study. J Clin Oncol. 2005;23(34):8646–54.

11. Bonner JA, Harari PM, Giralt J, Azarnia N, Shin DM, Cohen RB, et al. Radiotherapy plus cetuximab for squamous-cell carcinoma of the head and neck. N Engl J Med. 2006;354(6):567–78.

12. Vermorken JB, Mesia R, Rivera F, Remenar E, Kawecki A, Rottey S, et al. Platinum-based chemotherapy plus cetuximab in head and neck cancer. N Engl J Med. 2008;359(11):1116–27.

13. Hatakeyama H, Cheng H, Wirth P, Counsell A, Marcrom SR, Wood CB, et al. Regulation of heparin-binding EGF-like growth factor by miR-212 and acquired cetuximab-resistance in head and neck squamous cell carcinoma. PLoS One. 2010;5(9):e12702.

14. Donev IS, Wang W, Yamada T, Li Q, Takeuchi S, Matsumoto K, et al. Transient PI3K inhibition induces apoptosis and overcomes HGF-mediated resistance to EGFR-TKIs in EGFR mutant lung cancer. Clin Cancer Res. 2011;17(8):2260–9.

15. Hah JH, Zhao M, Pickering CR, Frederick MJ, Andrews GA, Jasser SA, et al. HRAS mutations and resistance to the epidermal growth factor receptor tyrosine kinase inhibitor erlotinib in head and neck squamous cell carcinoma cells. Head Neck. 2014;36(11):1547–54.

16. Heindl S, Eggenstein E, Keller S, Kneissl J, Keller G, Mutze K, et al. Relevance of MET activation and genetic alterations of KRAS and E-cadherin for cetuximab sensitivity of gastric cancer cell lines. J Cancer Res Clin Oncol. 2012;138(5):843–58.

17. Quesnelle KM, Grandis JR. Dual kinase inhibition of EGFR and HER2 overcomes resistance to cetuximab in a novel in vivo model of acquired cetuximab resistance. Clin Cancer Res. 2011;17(18):5935–44.

18. Wheeler DL, Huang S, Kruser TJ, Nechrebecki MM, Armstrong EA, Benavente S, et al. Mechanisms of acquired resistance to cetuximab: role of HER (ErbB) family members. Oncogene. 2008;27(28):3944–56.

19. Yonesaka K, Zejnullahu K, Okamoto I, Satoh T, Cappuzzo F, Souglakos J, et al. Activation of ERBB2 signaling causes resistance to the EGFR-directed therapeutic antibody cetuximab. Sci Transl Med. 2011;3(99):99ra86.

20. Wang X, Schneider A. HIF-2alpha-mediated activation of the epidermal growth factor receptor potentiates head and neck cancer cell migration in response to hypoxia. Carcinogenesis. 2010;31(7):1202–10.

21. Bedi A, Chang X, Noonan K, Pham V, Bedi R, Fertig EJ, et al. Inhibition of TGF-beta enhances the in vivo antitumor efficacy of EGF receptor-targeted therapy. Mol Cancer Ther. 2012;11(11):2429–39.

22. Levy EM, Sycz G, Arriaga JM, Barrio MM, von Euw EM, Morales SB, et al. Cetuximab-mediated cellular cytotoxicity is inhibited by HLA-E membrane expression in colon cancer cells. Innate Immun. 2009;15(2):91–100.

23. Brand TM, Iida M, Luthar N, Starr MM, Huppert EJ, Wheeler DL. Nuclear EGFR as a molecular target in cancer. Radiother Oncol. 2013;108(3):370–7.

24. Psyrri A, Yu Z, Weinberger PM, Sasaki C, Haffty B, Camp R, et al. Quantitative determination of nuclear and cytoplasmic epidermal growth factor receptor expression in oropharyngeal squamous cell cancer by using automated quantitative analysis. Clin Cancer Res. 2005;11(16):5856–62.

25. Hoshino M, Fukui H, Ono Y, Sekikawa A, Ichikawa K, Tomita S, et al. Nuclear expression of phosphorylated EGFR is associated with poor prognosis of patients with esophageal squamous cell carcinoma. Pathobiology. 2007;74(1):15–21.

26. Normanno N, Maiello MR, De Luca A. Epidermal growth factor receptor tyrosine kinase inhibitors (EGFR-TKIs): simple drugs with a complex mechanism of action? J Cell Physiol. 2003;194(1):13–9.

27. de Souza JA, Davis DW, Zhang Y, Khattri A, Seiwert TY, Aktolga S, et al. A phase II study of lapatinib in recurrent/metastatic squamous cell carcinoma of the head and neck. Clin Cancer Res. 2012;18(8):2336–43.

28. Kirby AM, A’Hern RP, D’Ambrosio C, Tanay M, Syrigos KN, Rogers SJ, et al. Gefitinib (ZD1839, Iressa) as palliative treatment in recurrent or metastatic head and neck cancer. Br J Cancer. 2006;94(5):631-6.

29. Soulieres D, Senzer NN, Vokes EE, Hidalgo M, Agarwala SS, Siu LL. Multicenter phase II study of erlotinib, an oral epidermal growth factor receptor tyrosine kinase inhibitor, in patients with recurrent or metastatic squamous cell cancer of the head and neck. J Clin Oncol. 2004;22(1):77–85.

30. Huang S, Armstrong EA, Benavente S, Chinnaiyan P, Harari PM. Dual-agent molecular targeting of the epidermal growth factor receptor (EGFR): combining anti-EGFR antibody with tyrosine kinase inhibitor. Cancer Res. 2004;64(15):5355–62.

31. Matar P, Rojo F, Cassia R, Moreno-Bueno G, Di Cosimo S, Tabernero J, et al. Combined epidermal growth factor receptor targeting with the tyrosine kinase inhibitor gefitinib (ZD1839) and the monoclonal antibody cetuximab (IMC-C225): superiority over single-agent receptor targeting. Clin Cancer Res. 2004;10(19):6487–501.

32. Kim HP, Yoon YK, Kim JW, Han SW, Hur HS, Park J, et al. Lapatinib, a dual EGFR and HER2 tyrosine kinase inhibitor, downregulates thymidylate synthase by inhibiting the nuclear translocation of EGFR and HER2. PLoS One. 2009;4(6):e5933.

33. Lin CC, Calvo E, Papadopoulos KP, Patnaik A, Sarantopoulos J, Mita AC, et al. Phase I study of cetuximab, erlotinib, and bevacizumab in patients with advanced solid tumors. Cancer Chemother Pharmacol. 2009;63(6):1065–71.

34. Camp RL, Chung GG, Rimm DL. Automated subcellular localization and quantification of protein expression in tissue microarrays. Nat Med. 2002;8(11):1323–7.

35. Suh CH, Park HS, Kim KW, Pyo J, Hatabu H, Nishino M. Pneumonitis in advanced non-small-cell lung cancer patients treated with EGFR tyrosine kinase inhibitor: Meta-analysis of 153 cohorts with 15,713 patients: Meta-analysis of incidence and risk factors of EGFR-TKI pneumonitis in NSCLC. Lung Cancer. 2018;123:60–9.

36. Bauman JE, Saba NF, Roe D, Bauman JR, Kaczmar J, Bhatia A, et al. Randomized Phase II Trial of Ficlatuzumab With or Without Cetuximab in Pan-Refractory, Recurrent/Metastatic Head and Neck Cancer. J Clin Oncol. 2023;41(22):3851–62.

37. Cohen EEW FJ, Daste A, et al, editor Clinical activity of MCLA-158 (petosemtamab), an IgG1 bispecific antibody targeting EGFR and LGR5, in advanced head and neck squamous cell carcinoma (HNSCC). AACR Annual Meeting; 2023; Orlando, FL.

38. Glenn J. Hanna JMK, Dan Paul Zandberg, Deborah J.L. Wong, Emrullah Yilmaz, Eric Jeffrey Sherman, Alberto Hernando-Calvo, Assuntina G. Sacco, Christine H. Chung, David Bohr, Ralf Reiners, Rachel Salazar, Elham Gharakhani, Sanela Bilic, and Jameel Muzaffar, editor Dose expansion results of the bifunctional EGFR/TGFβ inhibitor BCA101 with pembrolizumab in patients with recurrent, metastatic head and neck squamous cell carcinoma. ASCO Annual Meeting; 2023; Chicago, IL: JCO 41, 6005-6005(2023).

39. Han; Kwangmin Na; Young Taek Kim; Sungwoo Lee; Mi Ran Yun; Jae Hwan Kim; Youngseon Byeon; Young Seob Kim; Jii Bum Lee; Ji Yun Lee; Chang Gon Kim; Min Hee Hong; Kyoung-Ho Pyo; Joshua Curtin; Bharvin Patel; Isabelle Bergiers SMLC-BSS-sKDKS-HLSBSMYYJHMhKH, editor Abstract 5865: Combinatorial activity of amivantamab and pembrolizumab in head and neck squamous cell carcinoma and lung squamous cell carcinoma expressing wild-type EGFR and MET. AACR Annual Meeting; 2023; Orlando, FL: Cancer Res (2023) 83 (7_Supplement): 5865.

